# Trajectories of neurodegeneration and seed amplification biomarkers prior to disease onset in individuals at risk of prion disease

**DOI:** 10.1101/2022.10.30.22281644

**Authors:** Tze How Mok, Akin Nihat, Nour Majbour, Danielle Sequeira, Leah Holm-Mercer, Thomas Coysh, Lee Darwent, Mark Batchelor, Bradley R Groveman, Christina D Orrù, Andrew G Hughson, Amanda Heslegrave, Rhiannon Laban, Elena Veleva, Ross W Paterson, Ashvini Keshavan, Jonathan Schott, Imogen J Swift, Carolin Heller, Jonathan D Rohrer, Alexander Gerhard, Christopher Butler, James B Rowe, Mario Masellis, Miles Chapman, Michael P Lunn, Jan Bieschke, Graham Jackson, Henrik Zetterberg, Byron Caughey, Peter Rudge, John Collinge, Simon Mead

## Abstract

Human prion diseases are remarkable for long incubation times followed by typically rapid clinical decline. Seed amplification assays and neurodegeneration biofluid biomarkers are remarkably useful in the clinical phase, but their potential to predict clinical onset in healthy people remains unclear. This is relevant not only to the design of preventive strategies in those at-risk of prion diseases, but more broadly, because prion-like mechanisms are thought to underpin many neurodegenerative disorders. Here we report the accrual of a longitudinal biofluid resource in patients, controls and healthy people at-risk of prion diseases, to which ultrasensitive techniques such as real-time quaking-induced conversion (RT-QuIC), and single molecule array (Simoa) digital immunoassays were applied for preclinical biomarker discovery. We studied a total of 648 CSF and plasma samples, including importantly, 16 people who had samples taken when healthy but later developed inherited prion disease (IPD) (“converters,” range from 9.9 prior to, and 7.4 years after onset). A second generation (IQ-CSF) RT-QuIC assay was used to screen symptomatic IPD samples, followed by optimisation for other IPDs, before the entire collection of at-risk samples was screened using the most sensitive assay. Glial fibrillary acidic protein (GFAP), neurofilament light (NfL), tau and ubiquitin carboxy-terminal hydrolase L1 (UCH-L1) levels were measured in plasma and CSF. IQ-CSF RT-QuIC proved 100% sensitive and specific for sporadic Creutzfeldt-Jakob disease (sCJD), iatrogenic (iCJD) and familial CJD phenotypes, and subsequently detected seeding activity in four CSF samples from three *PRNP* E200K carriers in the presymptomatic phase, one of whom converted shortly after but the other two remain asymptomatic after two and three years of follow up. A bespoke HuPrP P102L RT-QuIC showed partial sensitivity for P102L disease and was positive in a CSF sample from an individual at risk of P102L IPD. No compatible RT-QuIC assay iterations were discovered for classical 6-OPRI, A117V and D178N, and these at-risk samples tested negative with bank vole RT-QuIC. Plasma GFAP and NfL, and CSF NfL levels emerged as proximity markers of neurodegeneration in slowly progressive forms of IPDs, with highly statistically significant differences in mean values segregating normal control (together with IPD > 2 years to onset) from IPD < 2 years to onset and symptomatic IPD cohorts. The trajectories of biomarker change appeared to correspond to expected fast and slow clinical phenotypes of progression in IPD with plasma GFAP changes preceding NfL changes. We propose patterns of preclinical biomarker changes in prion diseases based on the presence of clinical, seeding and neurodegeneration features.

## Introduction

Prion diseases are transmissible and inevitably fatal neurodegenerative conditions characterised by recruitment of host-encoded cellular prion protein (PrP) into disease-associated polymeric assemblies which propagate by elongation and fission(1). The observed range of clinical and pathological expressions in humans, however, is strikingly heterogenous despite the shared fundamental disease mechanism(2). Sporadic Creutzfeldt-Jakob disease (sCJD), the most prevalent form, accounts for roughly 85% of the incidence of human disease, typically presenting with the triad of rapidly progressive dementia, ataxia and/or myoclonus. Inherited prion disease (IPD) caused by autosomal dominant highly-penetrant mutations in the prion protein gene (*PRNP*) comprises 10-15% of the incidence but produces a wide spectrum of clinical syndromes including CJD (CJD), fatal familial insomnia (FFI), Gerstmann-Sträussler-Scheinker (GSS) disease, peripheral PrP systemic amyloidosis from truncation mutations, and long-duration dysexecutive-apraxic syndromes seen in octapeptide repeat insertions (OPRIs)(3, 4). Acquired prion disease has historically attracted considerable media, political and public health attention, despite being the rarest manifestation. Relevant exposures include bovine spongiform encephalopathy prions in the diet, use of blood and blood products for variant CJD(5); cadaver-sourced human growth hormone(6), neurosurgery, and lyophilised dura mater(7) in iatrogenic CJD, and at mortuary feasts in the Eastern Highlands Province of Papua New Guinea in kuru(8).

One of the most remarkable aspects of prion biology is the apparent long incubation phase between prion infection/exposure and disease onset, lasting up to five decades in kuru and cadaver-sourced human growth hormone related iatrogenic CJD (iCJD)(6, 9). Prions are transmissible to laboratory rodents by inoculation allowing for study of the sequence of prion infection, propagation and toxicity which forms two mechanistically distinct phases(10-12). Specifically, following inoculation, infectious prion titres rise exponentially to reach a plateau which continues for a considerable time until disease onset. Infectivity and toxicity are therefore uncoupled, with the length of the plateau being inversely proportional to PrP expression level. If this two-phase kinetics model is applicable to human disease, the clinically silent incubation phase marked by high prion titres hypothetically offers a window of opportunity for discovery of fluid biomarkers that predict proximity to onset. Moreover, if borne out in human, the two-phase kinetics model would provide a foundation for targeted prevention strategies in prion disease(13, 14).

Longitudinal study of defined populations with high lifetime risk of prion disease undoubtedly affords the best opportunity to elucidate the sequence of biomarker evolution during the presymptomatic phase in humans. For context, cadaver-sourced human growth hormone (c-hGH) was administered to 1,883 UK individuals between 1958 and 1985 of whom 80 have so far succumbed to iCJD(6, 15), while those currently at risk of IPD were estimated at 1,000 in the UK(16). Accrual of longitudinal biofluid sample resources from these studies not only allows repeated examinations for biomarker discovery, but also for ascertainment of rates of change as an even more sensitive predictor of disease onset(17). It is not feasible to adequately power clinical trials for candidate drugs in prion disease prevention for a simple clinical endpoint(18), but the characterisation of presymptomatic biomarkers could inform different strategies, enrichment in and learning from trials.

The advent of ultrasensitive real-time quaking-induced conversion (RT-QuIC) assays capable of detecting PrP-amyloid seeding down to the attogram (10^-18^ g) range in the last decade offers the potential to detect presymptomatic CSF PrP-amyloid seeding activity in at-risk individuals(19, 20). The assay exploits the ability of PrP-amyloid in tested samples to convert recombinant PrP (rPrP) monomers within a reaction mixture, and accelerates the process with cyclical bursts shaking and rest to amplify rPrP amyloid fibrils; alteration of thioflavin T emission spectrum from amyloid binding within the reaction mixture is then detected by a microplate reader in relative fluorescence units (rfu) over a certain threshold. Indeed, Orru et al. 2012 used RT-QuIC to demonstrate high levels of seeding activity in brains and CSF of hamsters experimentally inoculated with 263K prions in the clinically silent incubation period before disease onset, paralleling prion bioassays in the two-phase kinetics model(21); interestingly, Vallabh et al. 2020 identified presymptomatic RT-QuIC seeding activity in an single elderly carrier of the E200K mutation(22). CSF RT-QuIC assays in human prion disease to date have been honed primarily to detect sCJD and IPD E200K seeds to high sensitivity (>90%) and specificity (∼100%), far outstripping of its utility in other IPD disease syndromes(23-28). Nevertheless, assay developments along the way have identified key factors (incubation temperature, sodium dodecyl sulphate (SDS), Hofmeister salts, etc.) and novel seed-substrate compatibilities (truncated hamster (Ha90) and bank vole (BV) rPrPs) which may pave the way for optimising RT-QuIC for the more fastidious seed species in IPD(29-32).

Neurodegenerative biomarkers in prion disease are essentially downstream products of either neuronal injury, astrogliosis and inflammation, or other secondary disease pathologies. While none of them are strongly discriminatory between neurodegenerative diseases, particularly with cross-sectional values, the tracking of biomarker dynamics over time may segregate mutation carriers approaching disease onset from aging effects in normal controls. The introduction of digital immunoassay platforms revolutionised biomarker detection sensitivity, now down to single molecule resolution (e.g., Singe molecule array, Simoa), instead of relying solely on overall chemiluminescence intensity(33). Recently, our Unit demonstrated segregation plasma tau and neurofilament-light (NfL) levels between IPD mutation carriers from symptomatic IPD individuals, and more importantly showed rising levels in the two years prior to symptom onset in small numbers of converting individuals examined, through use of Simoa assays(34). Further advances in Simoa technology since then allows for multiplex arrays measuring up to four candidate biomarkers, limiting depletion of precious biofluid resources. The National Prion Monitoring Cohort (NPMC) study in the United Kingdom was well-placed to address this unmet need, having recruited at-risk individuals with contemporaneous acquisition of longitudinal clinical, neuroimaging, neuropsychometric and neurophysiological data, along with assembling an expansive blood and CSF biofluid archive since 2008. In this study, we marshalled the combined utility of disease-specific PrP-amyloid seed amplification assay (RT-QuIC) and ultrasensitive multiplexed Simoa digital immunoassay platform to characterise biomarker discovery and evolution in individuals at risk of prion disease.

## Materials and methods

### Ethical statement and study participants

Biofluid samples from all at-risk and symptomatic prion disease used in this study were drawn from individuals enrolled into the NPMC with written consent. Blood samples were routinely drawn at each scheduled assessment from 2008 onwards, while acquisition of CSF samples started in 2015 following an amendment to existing ethical approval for the NPMC through the Scotland A Research Ethics Committee. All studies here were approved by the local Research Ethics Committee of UCL Institute of Neurology and the National Hospital for Neurology and Neurosurgery (NHNN).

The NPMC enrolled eligible individuals from October 2008 onwards, encompassing those symptomatic of all forms of prion disease (sCJD, iCJD, vCJD and IPD), asymptomatic individuals of at risk of IPD (IPD-AR), iCJD (iCJD-AR) and vCJD, and healthy controls. The IPD-AR population includes confirmed asymptomatic carriers of pathogenic *PRNP* mutations, and untested blood relatives of those affected by, or known to carry, pathogenic *PRNP* mutations. The iCJD-AR population in the NPMC is chiefly composed of recipients of cadaver-sourced human growth hormone up to 1985. The schedule of assessments, and hence biofluid sampling intervals, were administered according to the stratum in which a participant falls, determined by the projected rate of disease progression(35), and by clinical need. At each assessment, research blood samples were also taken with written informed consent from willing friends or non-blood relatives as controls.

For Simoa biomarker comparison, healthy control (HC) CSF samples were sourced from the Young-Onset Alzheimer’s Disease spouses and non-blood relatives, the British 1946 Birth Cohort (Insight-46), CONFLUID cohorts (healthy controls with no cognitive concerns and Mini Mental State Examination scores > 27), and NPMC (single at-risk individual subsequently mutation-negative on predictive testing); HC plasma samples and data were sourced from NPMC internally (friends and non-blood relatives of patients) and from non-mutation carriers the Genetic Frontotemporal Dementia Initiative cohort (GENFI). For CSF RT-QuIC analyses, control samples were sourced from Institute of Neuroscience & Physiology at University of Gothenburg, NHNN Neuroimmunology Laboratory (NHNN-NiCL) and from NPMC (single at-risk individual subsequently mutation-negative on predictive testing).

### Proximity to clinical onset/conversion in IPD-AR individuals

Age at onset in IPD is highly variable (SD∼10 years), therefore many people who carry IPD mutations are healthy beyond their parental or average age of onset for each mutation. Consequently, we developed a new method to estimate the age of onset for IPD-AR, whereby each individual has an estimated age of onset in the future. This method approximates a cumulative normal distribution of risk for each mutation based on literature data, and sets estimated age of onset in the future equal to the accrual of 50% of an individual’s outstanding cumulative risk. Further details and an example are provided in the Supplementary Materials section (Supplementary 1a).

### NPMC biofluid sample processing

#### Blood

Whole blood samples collected in EDTA or Citrate destined for fractionation into plasma are centrifuged at 2000g for 10 minutes at room temperature (22°C), on arrival to laboratory. The supernatant (upper plasma phase) is then divided into aliquots of 0.5 – 2.0 mls in Nunc Cryovials, and then frozen at −80°C.

#### CSF

CSF samples are collected in two separate polypropylene tubes (Sarstedt 62.610.018), designated as CSF-R (for RT-QuIC) and CSF-N (for neurodegenerative markers). CSF-R is divided into aliquots of 0.5-1 ml in Nunc Cryovials after gentle mixing. CSF-N is centrifuged at 2200g for 10 mins at room temperature, and supernatant separated into aliquots of 0.5-1.0 ml in Nunc Cryovials. Both are then stored in −80°C freezers.

### rPrP expression and purification

Full length human (FL Hu rPrP; aa residues 23-231; accession M13899) and bank vole rPrP (FL BV rPrP; aa residues 23-231; accession AF367624), and truncated hamster (Ha90 rPrP; aa residues 90-231; accession K02234) and truncated bank vole rPrP (BV90 rPrP; aa residues 90-231; accession AF367624) were purified according to previously established methods(36, 37). The full length human P102L rPrP (FL Hu P102L rPrP; aa residues 23-231; accession M13899) construct contained his-tags, and as such was purified using through a different protocol (38) with some minor modifications. Further details are available in Supplementary Materials (Supplementary 1b).

### CSF RT-QuIC Analyses

The standard RT-QuIC reaction mix per well is composed of 10 mM buffer (sodium phosphate pH 7.4, or HEPES pH 7.4 or 8.0), 130 – 300 mM NaCl or NaI, 0.1 mg/ml rPrP (Hu, BV, Ha90, BV90 or Hu P102L), 10 μM Thioflavin T (ThT), 1 mM ethylenediaminetetraacetic acid tetrasodium salt (EDTA), and 0.001 or 0.002% SDS. Reactions were prepared in 96-well optical clear-bottomed plates (Nalgene Nunc International 265301). In each well, 80 or 85 μL of reaction mix was seeded with 20 or 15 μL of CSF respectively, bringing the final volume up to 100 μL per well.

Thereafter, the loaded plates were sealed (Thermo Scientific Nunc 232702) and incubated in BMG FLUOstar Omega Lite or POLARstar Omega microplate readers between 42-55°C, at double orbital shake/rest cycles of 60s/60s at 700 rpm. ThT fluorescence readings (excitation 450 ± 10 nm, emission 480 ± 10 nm; bottom read) were recorded at intervals of 45 mins. Each sample was tested in quadruplicate and classed as positive if the relative fluorescence units (RFU) in ≥ 2/4 wells exceed the 10% baseline-corrected threshold(39) within the corresponding time cut-off points. Samples initially resulting in 1/4 positive wells were retested, and if they remain 1/4 wells positive, were classed as ‘equivocal’. Time cut-off points were determined by incubation temperature i.e. 50 hrs for 42°C, 30 hrs for 50°C, and 24 hrs for 55°C(29).

### Endpoint quantitation of CSF seeding activity

CSF seeding doses were determined through endpoint quantitation of RT-QuIC PrP-amyloid seeding activity using the Spearman-Kärber method originally used in animal bioassay(37, 40). Each sample was serially diluted by one third using a single non-prion control CSF sample to reconstitute the total seeding volume per well to 20 μl. We define 50% seeding dose (SD_50_) as a unit of seeding activity or end-point sample dilution that yields positive responses in 50% (e.g. 2 of 4) RT-QuIC reaction wells according to the criteria above. The SD_50_ can be estimated from the results of a dilution series using LogSD_50_ = *x*_*p= 1*_ + 1/2*d* - *d∑p* where *x*_*p = 1*_ being the highest log_10_ dilution with 4/4 positive wells; d = log dilution factor; p = proportion positive at a given dose; *∑p* = the sum of values of *p* for *x*_*p = 1*_ and all higher dilutions. Adjustments can then be made to report SD_50_ per unit of neat sample, e.g. undiluted CSF. When a neat CSF sample (20 μl) yielded only 3/4 positive wells, the Spearman-Kärber method was not used, and instead, this sample was calculated to contain 1.5 SD_50_ (per 20 μl CSF) because, by definition, 1 SD_50_ gives 2/4 positive wells.

### N4PB biomarker measurement

Plasma and CSF glial fibrillary acidic protein (GFAP), NfL, tau and ubiquitin C-terminal hydrolase 1 (UCH-L1) were measured by Simoa using the N4PB kit on a HD-X Analyser (Quanterix), following the manufacturer’s protocol(33). In brief, samples were thawed and centrifuged at 10,000g for 5 minutes at room temperature (21°C) to precipitate any debris; subsequently, the samples were transferred to designated wells on the plates, diluted at 1:4 for plasma and 1:40 (or 1:100) for CSF with sample diluent, and bound to paramagnetic beads coated with capture antibodies specific for human GFAP, NfL, tau and UCH-L1. Longitudinal samples from a single patient where available, were analysed on the same plate. The biomarker-bound beads were then incubated with the respective biotinylated detection antibodies which in turn are conjugated to streptavidin-β-galactosidase complex which serves as a fluorescent tag. Hydrolysis of the complex of a resorufin β-D-galactopyranoside substrate results in a fluorescent signal proportional to the concentration of the respective biomarkers present. Measurements from each sample were with biomarker concentrations extrapolated from a standard curve, fitted to a 4-parameter logistic algorithm. Coefficients of variation (CVs) were determined using four internal quality control samples, and were < 20% and < 10% for intra-assay and inter-assay comparisons.

Additional previously measured plasma NfL and GFAP values from GENFI non-mutation carriers were also used to supplement healthy control data; five samples from this group were also analysed in our study to validate that the inter-assay CVs were < 15% for these two plasma markers.

### Data and statistical analyses

Similar to previous experience, the N4PB biomarker values including healthy controls (HC) here were positively skewed(34). Log_10_ transformation of GFAP, NfL, tau and UCH-L1 values reduced skewness across our sample cohorts, rendering them approximately normally distributed, allowing group-wise comparison of means using Single Factor ANOVA followed by pairwise t-tests. To address the known normal aging effects on at least GFAP, NfL and Tau levels(41-44), biomarker values for HC CSF and IPD-AR CSF greater than two years to predicted onset were normalised to age 60 (apart from UCH-L1 which did not demonstrate an age effect). Single-factor ANOVA followed by pairwise t-tests (assuming α = 0.05) were then applied to compare means of age-normalised values grouped by the respective cohorts – healthy controls (HC), IPD at-risk individuals more than two years to predicted/actual clinical onset (IPD-AR>2y), IPD at-risk individuals less than two years to predicted/actual clinical onset (IPD-AR<2y), symptomatic IPD individuals (IPD), sCJD/vCJD/iCJD individuals (CJD), and iCJD at risk individuals (iCJD-AR). Individual biomarker slopes were modelled using mixed effects models, with random effects of slopes and intercepts. Statistical analyses were carried out using GraphPad Prism (version 9.2.0) and STATA v15.1.

## RESULTS

The range of *PRNP* mutations seen in our combined IPD and IPD-AR biofluid cohorts in this study included 5-OPRI, 6-OPRI, P102L, P105S, A117V, P157X, D178N-129V, D178N-129M, Y163X and E200K. For CJD, this included samples from sCJD, iCJD (human growth hormone) and vCJD; the iCJD-AR cohort only included recipients of implicated batches of cadaver-sourced human growth hormone. We defined clinical conversion as the emergence of characteristic neurological symptoms and signs along with functional decline measurable by MRC Prion Disease Rating Scale scores, supported by the presence of mutation-specific investigation abnormalities e.g. DWI abnormalities in E200K, neurophysiological abnormalities in P102L, polysomnographic abnormalities in D178N-FFI, etc.

### RT-QuIC PrP-amyloid seeding assay CSF sample cohorts

From 2015 to 2021, 161 CSF samples were accrued for RT-QuIC analysis; IPD-AR samples account for the largest proportion (n = 61; individuals = 39), followed by IPD (n = 20; individuals = 20), sCJD/iCJD (n = 17; individuals = 17) and c-hGH iCJD-AR (n = 4; individuals = 3), which were tested against non-prion controls (n = 59; individuals = 59). Three pairs of samples exist from E200K, 6-OPRI and P102L converters, each with one sample before and after conversion. Baseline demographic details are summarised in Table 1. The entire at-risk and converter cohort is depicted graphically in Fig. 1.

**Table 1.** Baseline demographics of N4PB and RT-QuIC cohorts.

| Plasma N4PB |  |  |  |  |  |  |  |  |  |  |
| --- | --- | --- | --- | --- | --- | --- | --- | --- | --- | --- |
| Cohorts | Cohort Subgroups | Number of samples | Unique Individuals | Mean Age at Sample in yrs (SD) | Female | Male | c129 MM | c129 MV | c129 VV | PRNP untested/unknown |
| IPD-AR |  | 217 | 69 | 43.9 (13.3) | 128 | 89 | 96 | 57 | 0 | 64 |
|  | IPD-AR > 2 yrs | 198 | 66 | 49.0 (13.2) | 115 | 83 | 82 | 52 | 0 | 64 |
|  | IPD-AR < 2 yrs | 19 | 14 | 43.4 (13.3) | 13 | 6 | 14 | 5 | 0 | 0 |
|  | *P102L* | 100 | 33 | 43.7 (10.4) | 68 | 32 | 26 | 25 | 0 | 49 |
|  | *E200K* | 59 | 22 | 52.2 (15.9) | 23 | 36 | 44 | 7 | 0 | 8 |
|  | Miscellaneous IPD | 58 | 15 | 35.9 (9.17) | 37.0 | 21 | 31 | 25 | 0 | 2 |
| h-GH iCJD-AR |  | 3 | 2 | 55.7 (0.6) | 1 | 2 | 1 | 0 | 0 | 1 |
| Symptomatic IPD |  | 62 | 26 | 50.9 (11.8) | 40 | 22 | 33 | 29 | 0 | 0 |
| CJD |  | 40 | 18 | 52.0 (14.5) | 12 | 28 | 16 | 19 | 5 | 0 |
| Normal controls | GFAP & NfL | 132 | 84 | 49.7 (13.5) | 66 | 66 | 0 | 0 | 0 | 127 |
|  | Tau & UCH-L1 | 89 | 41 | 51.9 (13.1) | 38 | 51 | 0 | 0 | 0 | 89 |
| CSF N4PB |  |  |  |  |  |  |  |  |  |  |
| IPD-AR |  | 67 | 40 | 46.9 (12.4) | 36 | 31 | 29 | 15 | 0 | 21 |
|  | IPD-AR > 2 yrs | 64 | 37 | 47.0 (12.2) | 33 | 31 | 26 | 15 | 0 | 21 |
|  | IPD-AR < 2 yrs | 3 | 3 | 46.4 (19.2) | 3 | 0 | 3 | 0 | 0 | 0 |
|  | *P102L* | 30 | 16 | 46.0 (12.1) | 18 | 12 | 6 | 5 | 0 | 19 |
|  | *E200K* | 23 | 16 | 54.1 (10.7) | 11 | 12 | 16 | 5 | 0 | 2 |
|  | Miscellaneous IPD | 14 | 8 | 37.3 (8.4) | 7 | 7 | 7 | 5 | 0 | 2 |
| h-GH iCJD-AR |  | 5 | 4 | 53.8 (3.0) | 1 | 4 | 1 | 0 | 0 | 4 |
| Symptomatic IPD |  | 22 | 21 | 48.4 (13.6) | 14 | 8 | 14 | 8 | 0 | 0 |
| CJD |  | 17 | 17 | 60.6 (10.7) | 10 | 7 | 7 | 9 | 0 | 1 |
| Normal controls |  | 24 | 24 | 69.1 (6.8) | 12 | 12 | 0 | 1 | 0 | 23 |
| CSF RT-QuIC |  |  |  |  |  |  |  |  |  |  |
| IPD-AR |  | 61 | 39 | 46.5 (12.3) | 31 | 30 | 28 | 13 | 0 | 20 |
|  | IPD-AR > 2 yrs | 58 | 36 | 46.5 (12.1) | 28 | 30 | 25 | 13 | 0 | 20 |
|  | IPD-AR < 2 yrs | 3 | 3 | 46.4 (19.2) | 3 | 0 | 3 | 0 | 0 | 0 |
|  | *P102L* | 27 | 16 | 45.4 (11.7) | 17 | 10 | 6 | 5 | 0 | 16 |
|  | *E200K* | 22 | 16 | 53.5 (10.8) | 10 | 12 | 15 | 5 | 0 | 2 |
|  | Miscellaneous IPD | 12 | 7 | 36.2 (8.6) | 5 | 7 | 7 | 3 | 0 | 2 |
| h-GH iCJD-AR |  | 4 | 4 | 53.25 (3.1) | 1 | 3 | 1 | 0 | 0 | 3 |
| Symptomatic IPD |  | 20 | 20 | 48.9 (13.4) | 13 | 7 | 12 | 8 | 0 | 0 |
| CJD |  | 17 | 17 | 59.4 (10.5) | 10 | 7 | 7 | 10 | 0 | 0 |
| Non-prion controls |  | 59 | 59 | 65.4 (14.5) | 28 | 31 | 0 | 1 | 0 | 58 |

Standard deviation (SD); codon 129 (c129); yrs (years); methionine homozygous (MM); methionine-valine heterozygous (MV); valine homozygous (VV)

**Figure 1.**
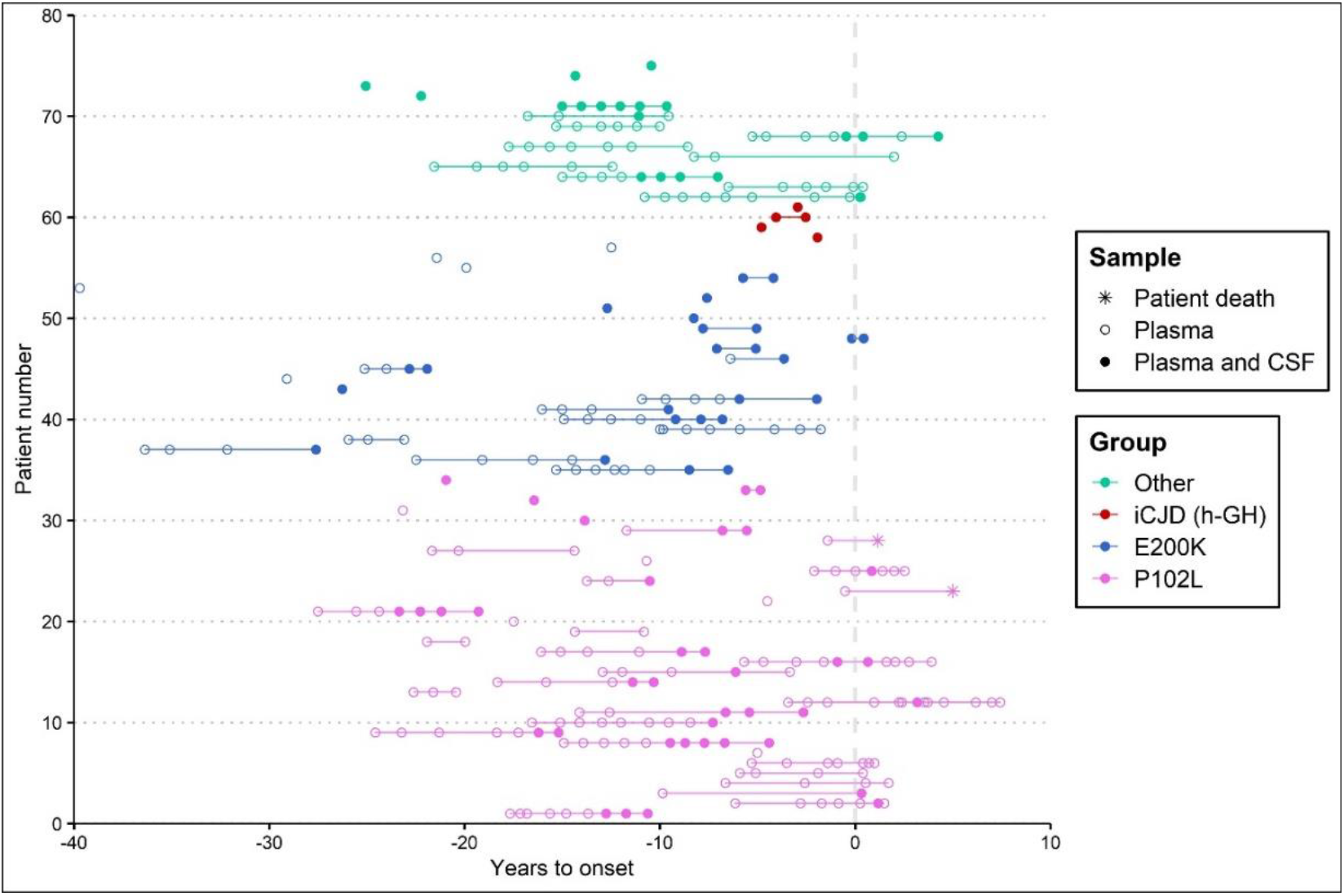
IPD-AR, iCJD-AR and IPD converter biofluid sample archive. This graph plots all the samples (plasma only versus paired plasma and CSF) analysed in this study against time to estimated or actual (known for converters) onset on the x-axis, marked by dotted vertical line at time 0. Samples from the same individual are joined by line if more than one sample was collected. For converters where only one presymptomatic sample exists without any follow-up samples, the subsequent data point joined by line indicates time of death. IPD mutations with fewer than five at-risk individuals were grouped as “Other” to avoid self-identification.

### N4PB neurodegenerative marker sample cohorts

We assembled a total of 416 plasma and 135 CSF samples, from 2008 to 2021, for Simoa PB measurements. The IPD-AR cohort again accounts for the majority of samples for both biofluids with 217 plasma samples from 69 unique individuals, and 67 CSF samples from 40 unique individuals. Crucially, this included longitudinal plasma (n = 86; individuals = 14) and CSF (n = 7; individuals = 3) samples which captured the interlude spanning clinical conversion (plasma range −9.9 to 7.4 years; CSF range −0.9 to 4.3 years); in 2 other IPD-AR individuals, a single plasma sample each was collected within 2 years of clinical conversion but none after. Of the 16 converted IPD individuals, 8 had plasma NfL and tau levels measured by simplex Simoa platforms and published previously(45). In addition to the IPD-AR cohort, plasma (n = 3; individuals = 2) and CSF (n = 5; individuals = 4) samples from asymptomatic h-GH iCJD-AR individuals were also tested against symptomatic IPD, CJD (sCJD, iCJD & vCJD), and HC cohorts, with baseline demographics summarised in Table 1.

### Optimisation RT-QuIC conditions for IPD CSF samples

A panel of CSF samples from clinically well-characterised individuals with symptomatic prion disease (IPD and CJD (sCJD and iCJD)) were first screened with IQ-CSF RT-QuIC (39). Subsequently, an exploratory set comprising of IQ-CSF RT-QuIC negative samples were put through iterative RT-QuIC assays with alterations in pH, buffer, incubation temperatures, salts, CSF seeding volumes, and rPrP species to determine the best available conditions for each IPD mutation, prior to testing the entire at-risk sample cohort.

Initial IQ-CSF RT-QuIC survey of the CJD sample set gave fifteen positive (≥ 2/4 wells) and two equivocal (1/4 wells) results, with the equivocal samples becoming positive after adjustment of seeding volume from 20 μl to 15 μl). All four CSF from symptomatic E200K carriers were strongly positive with IQ-CSF RT-QuIC where all wells from each became positive within 10 hrs of incubation. CSF from a single 6-OPRI carrier drawn following an unexpected ‘CJD-like’ transformation with corresponding DWI changes on MRI brain indistinguishable from sCJD after several years of classical 6-OPRI disease progression, was also strongly positive (Supplementary Fig. 2). CSF from symptomatic P102L, P105S, D178N-129M, Y163X and classical 6-OPRI were all negative. All control CSF samples (n = 59) tested negative for IQ-CSF RT-QuIC with 20 μl seeding volume, as did all the control CSF (n = 47) using 15 μL seeding volume. Overall, the IQ-CSF RT-QuIC sensitivity and specificity for CJD (sensitivity 95% CI (89.49, 100.00); specificity 95% CI (93.94, 100.00)) and E200K IPD (sensitivity 95% CI (39.76, 100.00); specificity 95% CI (93.94, 100.00)) samples were both 100%.

A new bespoke variation of RT-QuIC using rHuPrP P102L (PBS pH 7.4, 130 mM NaI, 0.002% SDS, 42°C) was positive in four of nine symptomatic P102L carriers. Of note, all four positive samples were from those with classical GSS phenotypes at onset, though one underwent a ‘CJD-like’ transformation featuring typical DWI MRI brain changes after two years(46). The samples from the remaining three symptomatic P102L individuals with purely cognitive phenotypes tested negative with Hu P102L, wild-type Hu, BV, IQ-CSF RT-QuICs and all other exploratory conditions.

CSF seeding activity (3/4 wells) in a symptomatic P105S carrier with a CJD-like phenotype with cortical ribboning on DWI MRI Brain, was best demonstrated using Hu rPrP in pH 7.4 with 130 mM NaI at 42°C (Supplementary Fig. 3).

Optimum RT-QuIC conditions for D178N-129M, Y163X and classical 6-OPRI were not found despite extensive exploration. In instances where seeding activity was demonstrated, they occurred beyond the cut-off time and frequently in close proximity to spontaneous fibrillisation in control wells.

### RT-QuIC analyses of IPD-AR and iCJD-AR CSF cohorts

We divided the at-risk samples into the following groups, and matched them to the best available RT-QuIC assay determined in the exploratory phase:

1. E200K-AR and iCJD-AR to Ha90 rPrP in pH 7.4 and 300 mM NaCl at 55°C (IQ-CSF RT-QuIC)
2. P102L-AR to Hu P102L rPrP in pH 7.4 and 130 mM NaI at 42°C (Hu P102L RT-QuIC) and BV rPrP in pH 7.4 and 300 mM NaCl at 50°C (BV RT-QuIC)
3. Other-AR to BV RT-QuIC

IQ-CSF RT-QuIC survey of the E200K-AR cohort (n = 22) revealed four positive results. All of these samples recorded 4/4 wells positive apart from one sample (8.3 years to onset) in which 3/4 wells were positive (Fig. 2A). CSF SD_50_/μl estimates were calculated as described in Methods. A pair from these samples belonged to an E200K converter, one 0.2 years before and the other 0.4 years after disease onset; the other pair was from an asymptomatic E200K carrier drawn at 7.1 and 5.1 years prior to estimated onset. The SD_50_/μl for the converter rose from 1.78 to 2.34, while that from the asymptomatic carrier dropped from 1.35 to 0.78 (Fig. 3).

**Figure 2.**
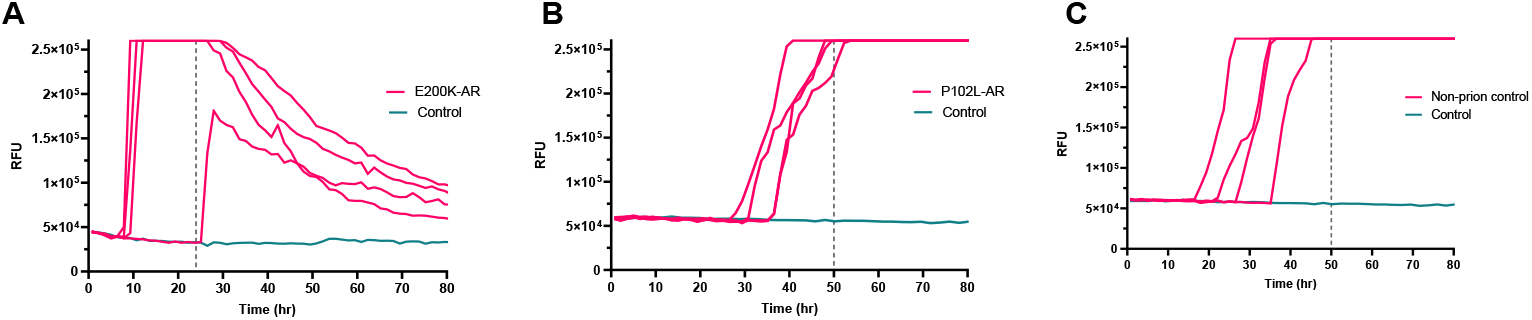
Graphs of select IPD-AR and control samples with positive RT-QuIC results. (**A**) This is the sole IQ-CSF RT-QuIC positive E200K-AR sample which recorded fewer than 4/4 wells positive, drawn at an estimated 8.3 years from disease onset. (**B**) This is the sole HuPrP P102L RT-QuIC positive sample in the P102L-AR set; this sample was negative when tested with BV RT-QuIC. (**C**) This non-prion disease (neurodegenerative) CSF sample tested positive with Hu P102L RT-QuIC, but tested negative with IQ-CSF RT-QuIC and BV RT-QuIC. The dotted vertical lines indicate the time cut-offs for the individual assays i.e. 24 hours for IQ-CSF RT-QuIC and 50 hours for Hu P102L RT-QuIC.

**Figure 3.**
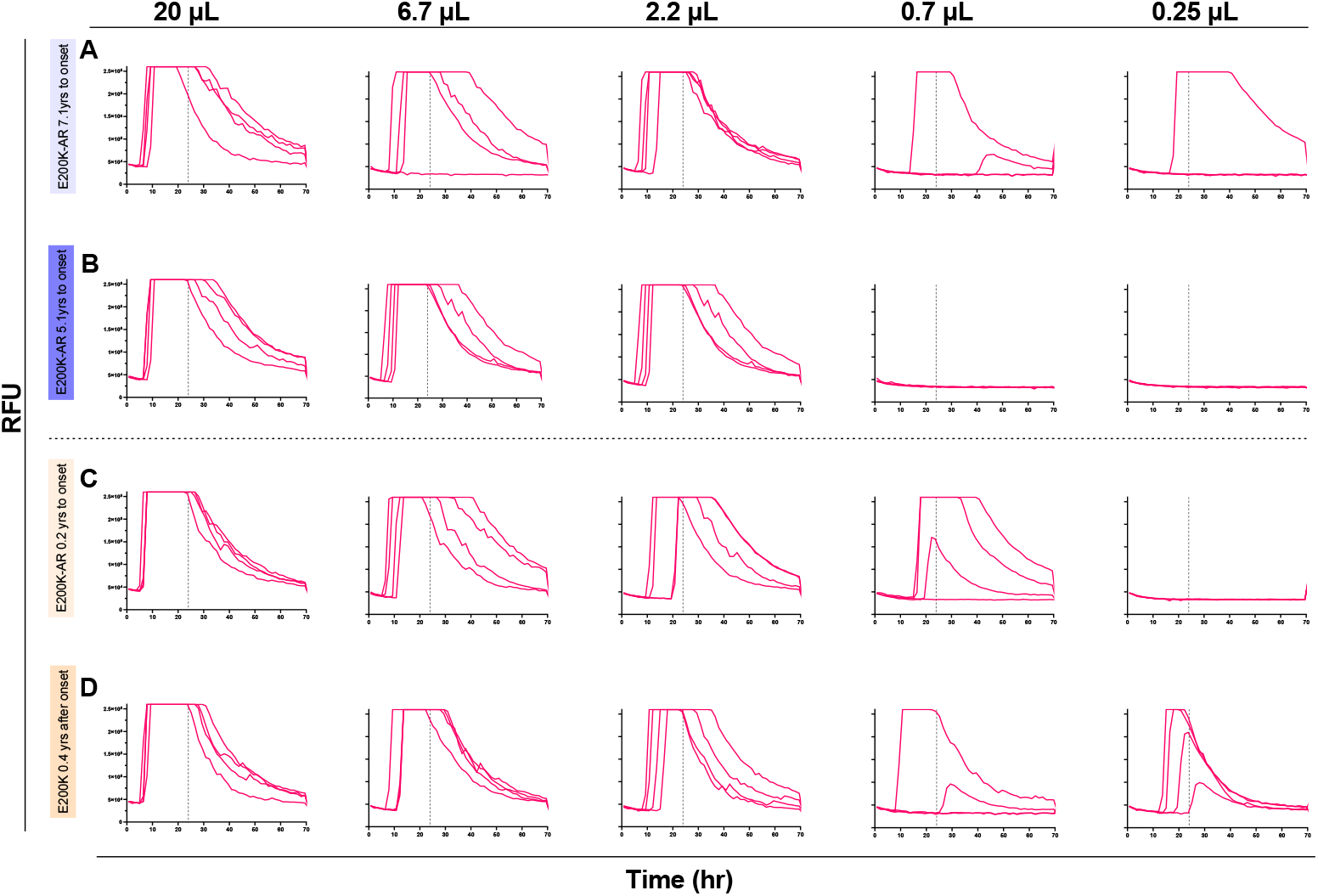
RT-QuIC CSF endpoint dilutions for E200K-AR and E200K converter samples to calculate SD50/μl. Each panel series show dilutions of seeding E200K CSF volume by a third; the vertical dotted line indicates the time cut-off which is 24 hours for IQ-CSF RT-QuIC. (**A**) and (**B**) are from a single individual two years apart, at 7.1 and 5.1 years to estimated conversion respectively. (**C**) and (**D**) are from a single converter individual 0.6 years apart at 0.2 years to, and 0.4 years after conversion respectively. The dotted vertical lines indicate the time cut-offs for the individual assays i.e. 24 hours.

In the P102L-AR subgroup first tested with Hu P102L RT-QuIC, all samples were negative apart from one (21/22) sample from an asymptomatic at-risk untested individual over the age of 60; 1/57 non-prion control also tested positive (Fig. 2B and Fig. 2C). Both these samples remained positive on repeat testing; of note, this non-prion control sample tested negative in both IQ-CSF and BV RT-QuIC assays. The sole P102L-AR sample linked to a clinical converter, drawn 0.9 years prior to disease onset was negative, but the sample drawn 0.6 years after clinical onset tested positive (3/4 wells) with the Hu P102L RT-QuIC assay.

The Other-AR samples were tested with the BV RT-QuIC assay on the basis of BV rPrP being a potential “universal acceptor” in experiments seeded by brain homogenates(30). None of the Other-AR samples (0/12) nor in the P102L-AR subgroup (0/27) tested positive with BV RT-QuIC. All the non-prion control CSF samples were negative (n = 51).

### Plasma Simoa N4PB results

Of the four biomarkers tested, log(GFAP) and log(NfL) demonstrated sequentially incremental and statistically significant mean values between IPD-AR>2yrs and IPD-AR<2yrs, IPD and CJD cohorts on singe-factor ANOVA with post hoc groupwise comparisons (Fig. 4A; log(GFAP), IPD-AR>2yrs versus IPD-AR<2yrs p=0.0006, IPD-AR<2yrs versus healthy controls p=0.0004, IPD-AR<2yrs versus IPD p=0.0003; for log(NfL), IPD-AR>2yrs versus IPD-AR<2yrs p=0.002, IPD-AR<2yrs versus healthy controls p=0.002, and IPD-AR<2yrs versus IPD p=3.7×10^−6^). Of note, there were no significant differences in the mean values between the healthy control (HC) and IPD-AR>2yrs cohorts (*p* = 0.623 for log(GFAP); *p* = 0.298 for log(NfL)). The mean values of the N4PB biomarkers according to cohort divisions, and the *p* values from the single-factor ANOVA analyses are summarised in Table 2.

**Table 2.**
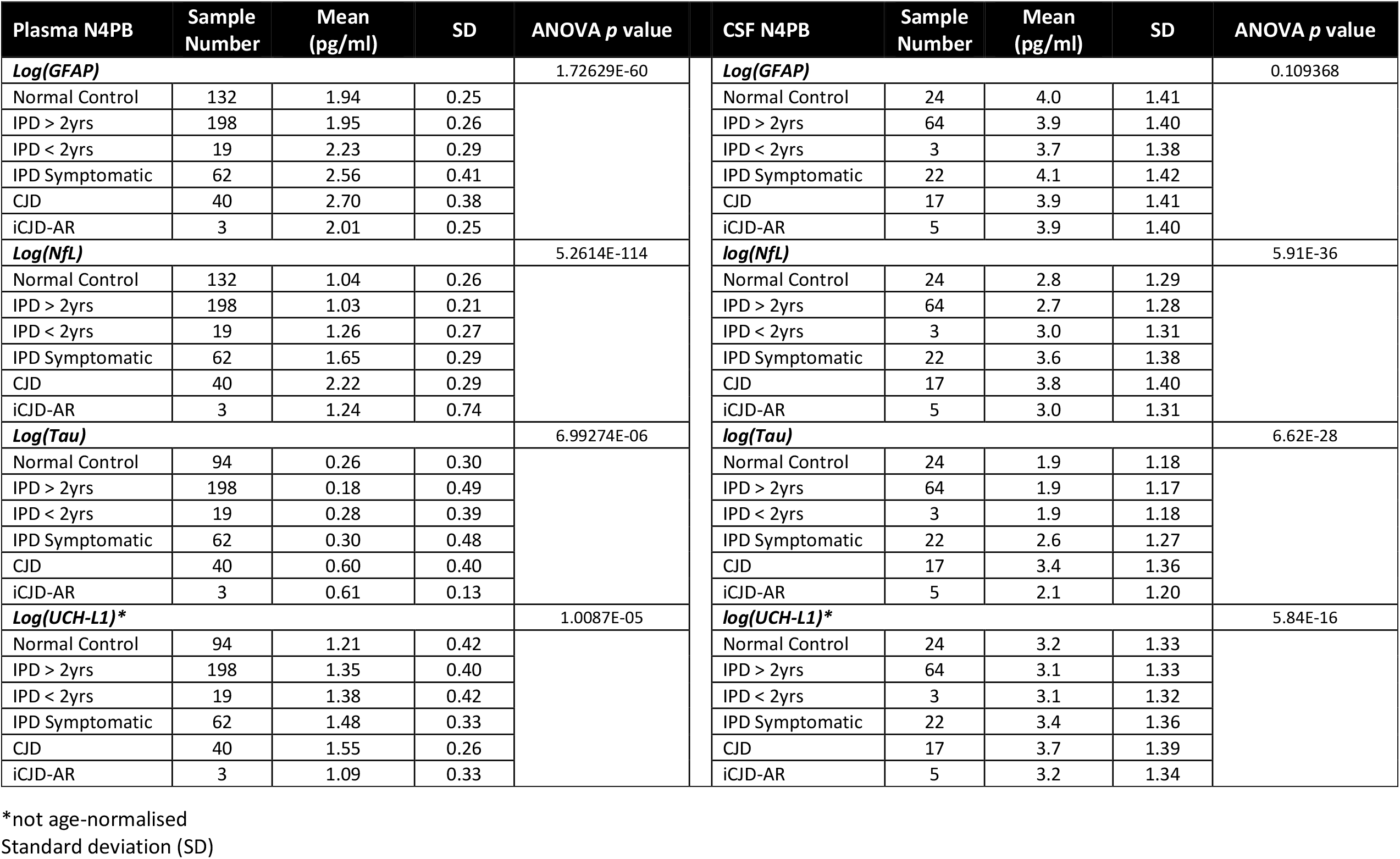
Mean values of age-normalised N4PB biomarkers according to cohort.

**Figure 4.**
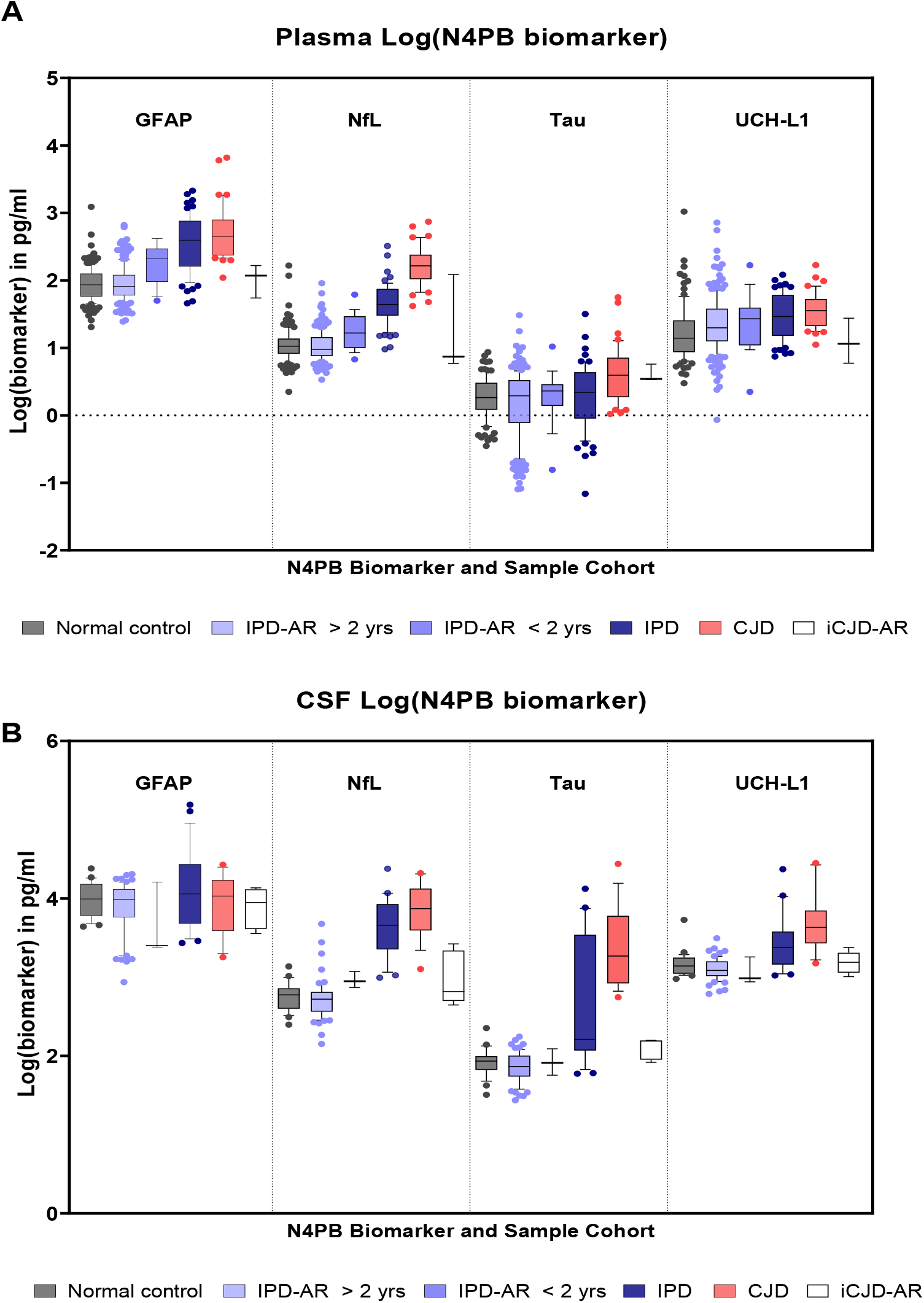
Simoa N4PB measurements in prion disease and at-risk cohorts. (**A**) Plasma N4PB levels, where only GFAP and NfL showed statistically significant different mean values between IPD at-risk and disease groups. (**B**) CSF N4PB levels, where only NfL showed statistically significant different mean values between IPD at-risk and disease groups.

Mean age-normalised log(Tau) was not statistically significant between IPD-AR>2yrs versus IPD-AR<2yrs (p = 0.329), Normal Control/IPD-AR>2 yrs versus IPD (p = 0.1), and IPD-AR<2 yrs versus IPD (p = 0.849). Mean log(UCH-L1) (not age-normalised) was not statistically different between IPD-AR>2yrs versus IPD<2yrs (p = 0.802). As for the iCJD-AR cohorts, relevant statistically significant mean values were only seen with log(GFAP) against the CJD cohort (2.01 vs 2.70 pg/ml; p = 0.02), and with log(Tau) against Normal Controls (0.609 vs 0.264 pg/ml; p = 0.03); the latter was driven by a single outlier in the sample obtained from an iCJD-AR individual with contemporaneous destructive pituitary craniopharyngioma.

We identified 16 *PRNP* mutation carriers (P102L = 10, D178N-FFI = 2, E200K = 1, 5-OPRI = 1, 6-OPRI = 1) who underwent clinical conversion during follow up over a median of 7.8 years (interquartile range 5.2 years) in whom at least one presymptomatic plasma sample was available for analysis. An incline in plasma log(GFAP) and log(NfL) values was observed, but most consistently in P102L, D178N-FFI and E200K converting individuals. The pattern of log(GFAP) and log(NfL) evolution for the clinically *fast IPD* converters (D178N-FFI and E200K) tend to exhibit relatively flat lines followed by abrupt rises close to or at the time of clinical onset. In comparison, the clinically *slow IPD* converters (P102L) showed a slower but more consistent upward trajectory in log(GFAP) and log(NfL) values, with 52.6% (10/18) and 44.4% (8/18) of measurements above the 90^th^ percentile of healthy controls (HC90) respectively in the 2 years before clinical onset (Fig.5A and Fig. 5B). None of the iCJD-AR individuals had converted to iCJD on follow up, but one died of invasive craniopharyngioma and another developed early onset Alzheimer’s dementia.

**Figure 5.**
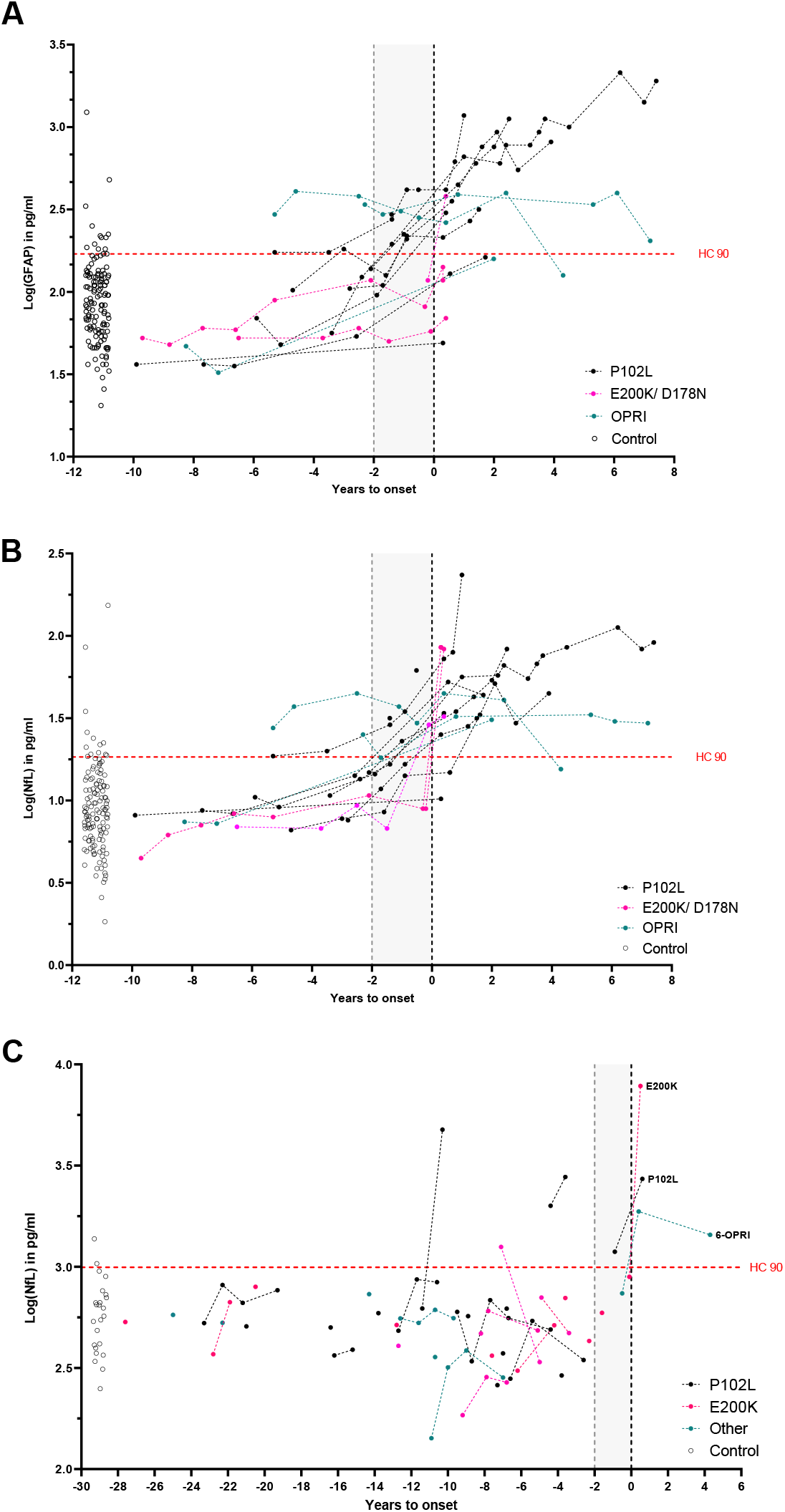
Converter trajectories for plasma GFAP and NfL, and CSF NfL. Plasma GFAP (**A**) and NfL (**B**) trajectories are grouped into P102L (slow IPD archetype) and E200K + D178N-FFI (fast IPD archetype) and OPRIs (slow IPD; includes 5- and 6-OPRI). Given the paucity of CSF converter data points, the entire CSF at-risk and converter cohort is shown in (**C**). The horizontal dotted line indicates the 90^th^ percentile of the respective biomarker value in the normal control cohort.

We therefore modelled the linear trajectories of presymptomatic NfL and GFAP using mixed effects regression models with random effects for individual slopes and “*fast* IPD” or “*slow* IPD” as factor variables, including data prior to conversion (up to four years prior to conversion for *slow* IPD; six months for *fast* IPD) and one time point up to six months after conversion. These models estimated a slope for plasma log(NfL) of 0.108 pg/ml/year in *slow* IPD (95% CI 0.0662-0.149) and 1.279 pg/ml/year in *fast* IPD (1.006-1.551) with an *x*-intercept (time pre-conversion that linear modelled trajectory crosses mean of controls) of 2.448 years; for plasma log(GFAP) 0.090 pg/ml/year in *slow* IPD (95% CI 0.040-0.140) and 0.458 pg/ml/year in *fast* IPD (0.129-0.787) with a *x*-intercept of 4.009 years.

### CSF Simoa N4PB results

In the CSF cohort, only log(NfL) successfully demonstrated incremental, and statistically significant segregation of the mean values between the IPD>2yrs, IPD<2yrs and IPD stages (Fig. 4B; for IPD-AR>2yrs versus IPD-AR<2yrs p=0.03, IPD-AR<2yrs versus healthy controls p=0.04, and IPD-AR<2yrs versus IPD p=2.95×10^−5^. No significant differences were shown between disease stages with log(GFAP). Statistically significant differences were seen for log(Tau) and log(UCH-L1) between IPD>2 yrs and IPD<2yrs versus IPD, but not between IPD>2 yrs versus IPD<2yrs.

In the three asymptomatic *PRNP* mutation carriers who clinically converted on follow up, only one individual (P102L) registered a CSF log(NfL) value (3.07 pg/ml) above the HC90 (2.97 pg/ml) in the two years prior to clinical onset; all the other N4PB biomarkers in the remaining two converters were below the HC90 values (Fig. 5C). All three converters exhibited overall rises in N4PB biomarker levels after conversion, but not all exceeded the HC90 threshold. Correspondingly, in the matched plasma sample drawn at the same time as the CSF samples, all N4PB biomarker levels were below the HC90 for the E200K converter 0.2 yrs before clinical onset; for the 6-OPRI converter, both plasma log(GFAP) and log(NfL) were above the HC90 at 0.5 years prior to onset, while for the P102L converter, only plasma log(GFAP) exceeded the HC90 at 0.9 years prior to onset.

## DISCUSSION

This study features extensive biomarker development in a large biofluid archive from individuals at risk of prion disease or in the early symptomatic stages. These data provide evidence for two types of fluid biomarker trajectory prior to clinical onset. Firstly, linear increases initially in GFAP, and later NfL, in *slow* forms of IPD up to four years pre-conversion. In contrast, in *fast* IPDs (CJD and FFI phenotypes), the neurodegeneration biomarkers (NfL) change explosively around onset with no definable presymptomatic window. Secondly, in those IPDs for which we have highly sensitive seed amplification assays, particularly those caused by the E200K mutation typically associated with a CJD-like phenotype, we found evidence of a potentially long duration presymptomatic CSF RT-QuIC seeding stage. These distinct aspects of prion pathophysiology are consistent with the two-phase kinetics model of prion propagation(10). They allowed us to envisage methods that might be used to stratify at-risk individuals and help the design and interpretation of presymptomatic treatment trials. It is important to note that that *bona fide* prion infectivity measured by two-phase kinetics(10) may be distinct from PrP-amyloid seeding by RT-QuIC, in that RT-QuIC can be seeded by non-infectious aggregated PrP.

Our analytical approach was underpinned by the expectation that any RT-QuIC assay ought to be sufficiently sensitive for detecting seeding activity in symptomatic CSF samples in order to be able to do so in presymptomatic samples. The compatibility of IQ-CSF RT-QuIC for CJD and E200K seeds was confirmed in the symptomatic cohort, and then in the at-risk sample set by picking up four positive samples, one of which belonged to a subsequent converter. This argues that the previously reported asymptomatic positive RT-QuIC in an E200K carrier was not an isolated finding(22). In our Cohort, no E200K at-risk individual converted without presymptomatic seeding activity. The E200K presymptomatic seeding period (as early as 8.3 years before predicted onset) appears unexpectedly long for an illness with such an explosive onset and short duration. None of the presymptomatic RT-QuIC positive samples, including one drawn shortly before conversion, recorded abnormal neurodegeneration biomarkers, indicating that the onset of neurodegeneration is likely to be very close to conversion and potentially unrecognisable at current sampling intervals.

CSF from P102L affected individuals has historically been tested by variations of PQ-CSF and IQ-CSF RT-QuIC, usually included as very small subsets within large surveys of national CJD surveillance cohorts, with low sensitivities(24-27, 47, 48) (Sano et al. 2013(28) is an exception). P102L individuals in these papers were classified as GSS with little information provided about clinical phenotype. Given the recognised phenotypic heterogeneity (classical GSS, cognitive & CJD-like) of P102L disease, and molecular evidence that these may be driven by distinct prion strains and possibly by non-infectious PrP amyloids accumulation, it is difficult to compare the results. It is quite possible that the few RT-QuIC positive samples reported may very well be due to the enrichment of individuals with the CJD-like clinical phenotype within surveillance cohorts(49-51).

We developed a bespoke RT-QuIC assay using Hu P102L rPrP and NaI capable of detecting CSF seeding activity in a subset of P102L diseased individuals and a single untested at-risk individual over 60 years of age (6.1 years to estimated onset; all relevant N4PB values < HC90). Detailed phenotypic profiling suggest that this assay may work best in the P102L-GSS and P102L-CJD subgroups, but not in the P102L-Cognitive subgroup but due to the small numbers tested, it is unknown whether this observation will hold true when applied to larger P102L CSF sample sets. It is possible that the one RT-QuIC positive control sample belonged to an undiagnosed P102L patient, as it was sourced from a cohort of patients with neurodegenerative symptoms, subsequently classed as either Alzheimer’s or non-Alzheimer’s disease based on CSF biomarker. Despite its partial sensitivity, our assay may have a role in identifying a subset of at-risk individuals whose future conversion will be driven by compatible P102L PrP isoforms. As for neurodegenerative markers, plasma log(GFAP) and log(NfL) trajectories together with a considerable proportion (including CSF log(NfL)) being above HC90 in the 2 years before onset denote a longer pre-conversion phase of escalating toxicity relative to E200K. *Slow* IPDs may possess an extended pre-conversion seeding window if appropriately sensitive assays can be developed, but more clearly, show an identifiable presymptomatic neurodegeneration window.

The promise of BV rPrP as a “universal acceptor” did not materialise during the RT-QuIC optimisation phase, despite efforts to improve sensitivity. No presymptomatic CSF seeding activity was detected in classical 6-OPRI, P102L, A117V, D178N-129M and D178N-129V at-risk samples using BV RT-QuIC despite previous demonstrations that brain homogenates (10^−4^ dilutions) of all but D178N-129V cases can be detected using BV RT-QuIC(30). Neither plasma log(NfL) nor log(GFAP) appeared helpful in identifying D178N-129M (FFI considered a *fast IPD*) individuals at risk of incipient conversion as the neurodegenerative window is as short as E200K. Little conclusion can be drawn as yet from the inconsistent N4PB biomarker trajectories for our small number of 5-OPRI and 6-OPRI converters.

We propose a general outline of presymptomatic biomarker change featuring key aspects of seeding activity, neurodegeneration, and clinical elements (Fig.6). Broadly, we saw patterns that vary for seeding and neurodegeneration between *fast* and *slow IPDs*. In *fast* IPD there is no useful presymptomatic neurodegeneration window at sampling intervals feasible in our study (Fig.6). Neurodegeneration trajectories for *slow* IPD are easy to discern in retrospect, however we cannot yet be confident enough for individual prediction in isolation as values lie within the range of healthy controls. Accurate prediction for the purposes of individual feedback is self-evidently essential given that the information is so consequential. Counterintuitively, plasma biomarker dynamics appear to hold more promise that CSF, but this may be artefactual, merely reflecting the relative lack of sampling and follow-up data points in the latter. The more immediate use may be for clinical trials, where we envisage the potential for biomarker-based enrichment of recruitment and biomarker outcomes in presymptomatic IPD. We believe that international collaboration will be essential to develop sample collections with sufficient power to build confidence in these patterns of change.

**Figure 6.**
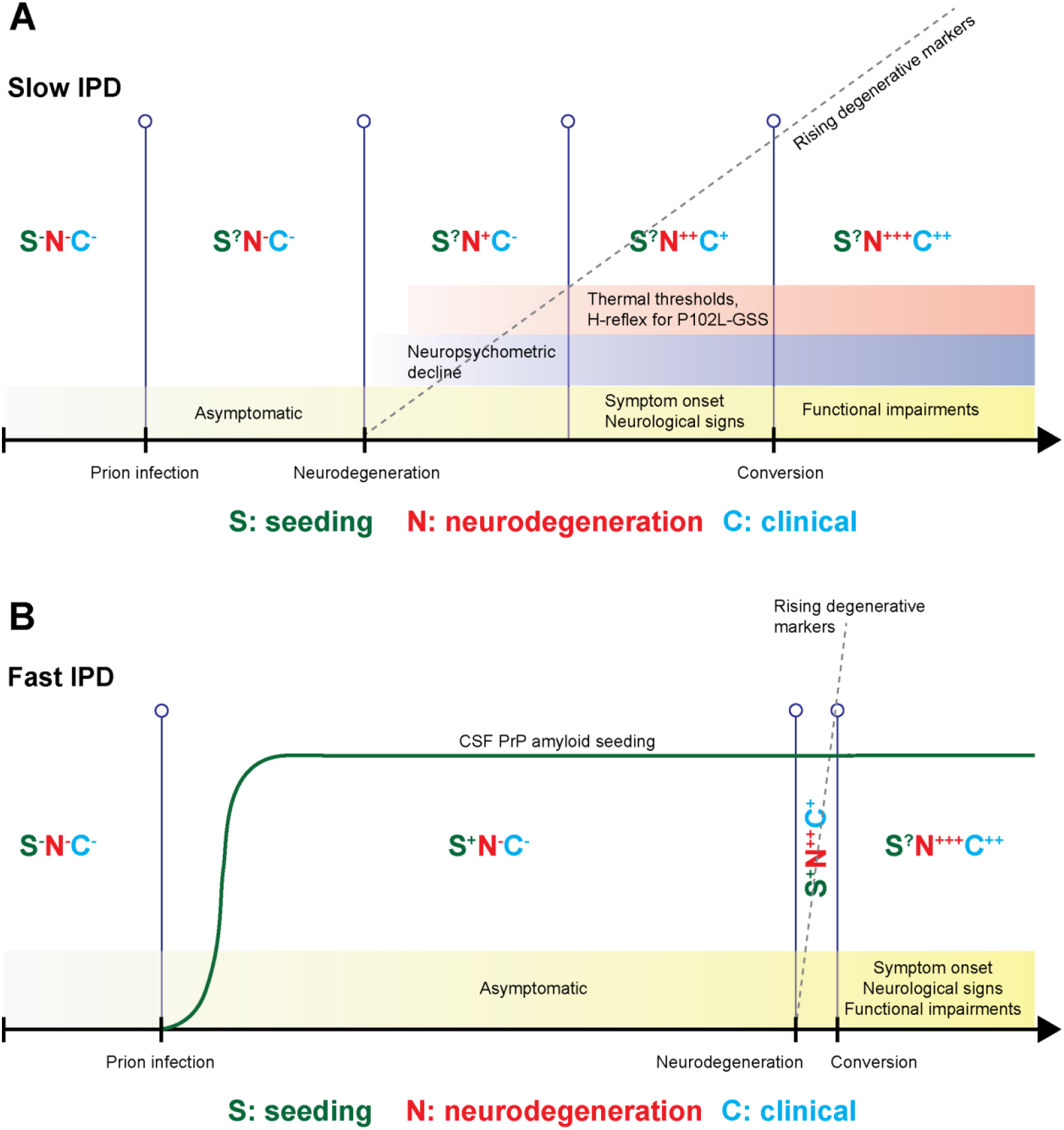
Proposed pre-conversion IPD patterns of biomarker change for fast and slow IPDs. Each stage features graded intensities in PrP-amyloid seeding activity, neurodegeneration markers and clinical aspects, along with ancillary investigations known to herald the onset of conversion (neuropsychometry, and neurophysiology in P102L). (**A**) Slow IPDs are likely to have an extended window for neurodegenerative markers, making it easier to capture and follow at 6-12 monthly sampling intervals; however, we only have partially sensitive RT-QuIC seeding assays for slow IPDs. (**B**) Fast IPDs are likely to have a very short and explosive neurodegeneration window, which means it might not be easy to capture and follow at similar sampling intervals; this may be offset by the existence of highly sensitive RT-QuIC assays that may become positive several years before clinical onset. The changes in CSF PrP amyloid seeding are hypothetical, current evidence is limited to a very small number of individuals and samples.

If seeding assays can be more widely developed and applied in neurodegenerative diseases, as seems likely, these findings might provoke exploration of long presymptomatic seeding phases in other disorders. Discoveries in recent years have revealed fundamental aspects of common neurodegenerative diseases similar to prion diseases, particularly in proteopathic seed propagation, transmissibility and strain biology(52-56). The RT-QuIC-type proteopathic seed amplification assay borne out of the prion disease field has the potential to extend the presymptomatic stage earlier than the neurodegenerative phase, which is already very well characterised by imaging, neuropsychometric, fluid markers, etc. in AD and FTD(57-59). Indeed, the adaptation of RT-QuIC for α-synuclein has been used to probe the pre-motor phase of Parkinson’s disease, Lewy body dementia and multiple systems atrophy with success(60-62). Furthermore, RT-QuIC for 3-repeat, 4-repeat and AD tau, and even TDP-43 are being honed for wider application is tissues and CSF(63-66). If successful, the extension of the presymptomatic phase to include a proteopathic seeding**-**only phase without evidence of neurodegeneration, will open even earlier windows of opportunity for intervention, with particular implications on timing, and of course study design for therapeutic strategies.

## Data Availability

All data produced in the present study are available upon reasonable request to the authors

## Acknowledgements

We thank all NPMC participants and their relatives for their unflinching dedication to this study, by volunteering their precious time and donating biofluid samples repeatedly even during times of debilitating illness and personal grief, for more than a decade. We thank Sarah Mazdon, and her predecessor Joanna Field, for managing and administrating the Cohort visits and investigations. We thank Richard Newton for creating and formatting the images for publication.

## Supplementary material

Supplementary material is available at *Brain* online.

## Supplementary Materials

### 1. Materials and Methods

#### a. Calculation of estimated time to disease onset

Assuming a normal distribution of age-related *z* scores against the probability density of clinical conversion, the *z* score associated with the current age (*z*_*c*_) of the individual is first determined, from which the cumulative distribution function at *z*_*c*_ (*P (z* < *z*_*c*_)) is derived. We then assume that the individual’s residual cumulative risk (area under the curve) lies to the right of z_c_ which is the inverse i.e (1 – *P (z* < *z*_*c*_)) and that the cumulative risk at *z*_*p*_ (*P (z* < *z*_*p*_)) is the sum of *P (z* < *z*_*c*_) and half of its inverse (1 – *P (z* < *z*_*c*_))/2 i.e., *P (z* < *z*_*p*_) = *P (z* < *z*_*c*_) + (1 – (*P (z* < *z*_*c*_)/2)). This can be expressed as a Microsoft Excel formula *x*_*p*_=NORM.INV(1-(1-NORM.DIST(A2,A3,A4,TRUE))/2,A3,A4)-A2 where A2 is the individuals current age (*x*_*c*_), A3 is the mutation mean age of onset, and A4 is the mutation standard deviation.

For example, the *z* score (*z*_*c*_) of an E200K IPD-AR individual aged 60 (*x*_*c*_) with a mutation mean (μ) of 58.5 and standard deviation (σ) of 8.0 years is calculated as follows:

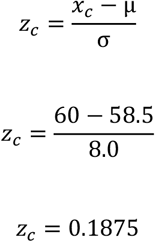

*P (z* < *z*_*c*_) of *z*_*c*_ (0.1875) is 0.5753, and therefore the inverse is 1 – 0.5753 = 0.4247. Half of 0.4247 is 0.4247/2 = 0.2123; and as such the *z*_*p*_ corresponds to *P (z* < *z*_*p*_) = 0.5753 + 0.2123 = 0.7876 is 0.8. Hence,

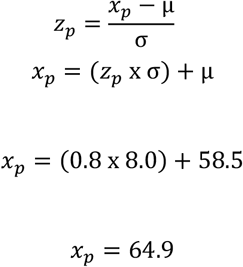

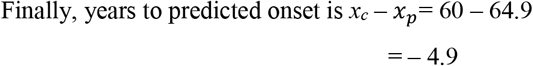

**Supplementary Figure 1.**
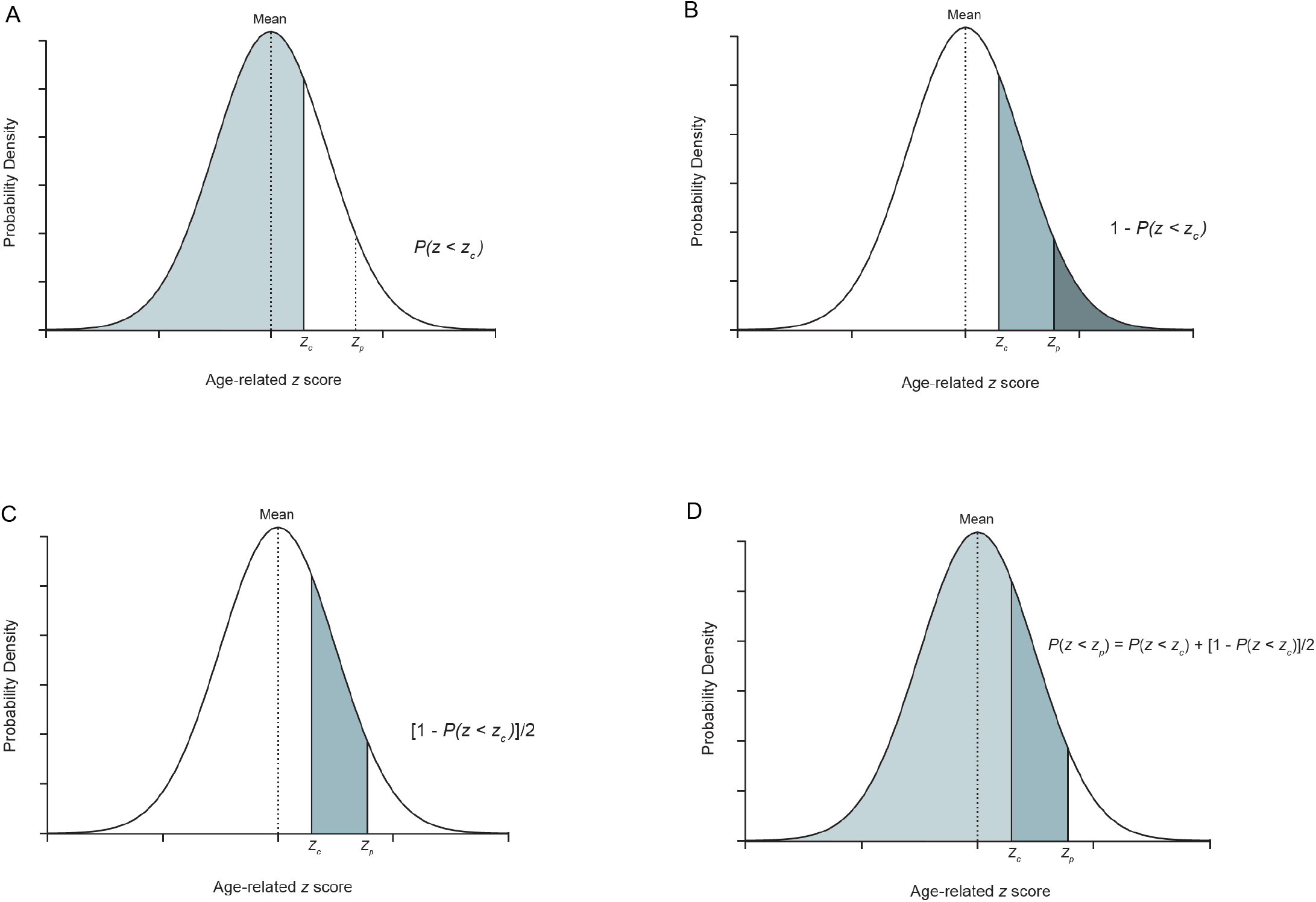
Determination of estimated age to onset for individuals at risk of prion disease (IPD and iCJD). (**A**) This shows the cumulative distribution function associated with the *z* score at the age of sampling (*P (z* < *z*_*c*_)). (**B**) The total shaded area is the cumulative distribution function of the residual risk (1 – *P (z* < *z*_*c*_)) and half of it is the estimated residual risk (**C**) for the individual at age of sampling (1 – (*P (z* < *z*_*c*_)/2)). So (**D**) represents the cumulative distribution function of the age-related *z* score at estimated onset *z*_*p*_ i.e. *P (z* < *z*_*c*_) + (1 – *(P (z* < *z*_*c*_)/2)).

#### b. Recombinant PrP expression and Purification

For Ha90, Hu, BV and BV90 rPrPs, glycerol stocks of Escherichia Coli with vectors containing the respective *PRNP* sequences above (sourced from the NIH Rocky Mountain Laboratory) were used to inoculate cultures which were subsequently grown in Luria Broth medium together with kanamycin and chloramphenicol. The autoinduction system was then used to stimulate protein expression. Purification of rPrP from inclusion bodies in denaturing conditions was done through a Ni-nitrilotriacetic acid (NiNTA) superflow resin (Qiagen) with an ÄKTA Pure, before refolding through a guanidine HCl gradient and elution through an imidazole gradient. The eluted rPrP was sequentially dialysed extensively in 10 mM of sodium phosphate buffer pH 5.8, filtered through 0.22 μm syringe filter, its concentration determination by absorbance measurement at 280 nm, separated into aliquots, and frozen at − 80°C. Prior to use, rPrP was thawed, filtered 100 kDa spin filter (Pall Nanosep), and concentration again similarly measured. The rPrP constructs purified with this method do not contain histidine tags (his-tags).

For Hu P102L rPrP, Escherichia Coli cultures containing the vector with this FL Hu P102L PrP sequence were grown Luria Broth medium, and in the presence of ampicillin. PrP expression was induced using Isopropyl β-D-1-thiogalactopyranoside (IPTG), and purified from inclusion bodies, similarly under denaturing conditions through NiNTA superflow resin (Qiagen) with an ÄKTA Pure, before refolding through a guanidine HCl gradient and elution through an imidazole gradient. The eluted rPrP was 1^st^ dialysed extensively against 20 mM Bis Tris pH 6.5, and then had its his-tags cleaved by addition of 2.5 mM CaCl_2_ and 50U Thrombin (VWR). The his-tags were when removed from the preparation by a 2^nd^ run through NiNTA superflow resin (Qiagen) with an ÄKTA Pure, dialysed against 10 mM of sodium phosphate buffer pH 5.8, and treated exactly as above at its corresponding stage of handling.

### 2. RESULTS AND DISCUSSION

#### a. 6-OPRI case with positive IQ-CSF RT-QuIC

The abrupt change in clinical course of the 6-OPRI affected individual whose late-stage CSF tested strongly positive with IQ-CSF RT-QuIC, raises the possibility of a “strain switch”. The initial MRI Brain showed only atrophy, with CJD-like DWI changes emerging only after onset of rapid decline. We postulate that somehow, misfolded wild-type PrP became the dominant isoform, with the earlier phase of the illness being driven by classical 6-OPRI PrP isoforms. After all, 6-OPRI disease is known to present like sCJD from the outset and there are a handful of cases from the literature featuring less than a year’s disease duration (67, 68). These cases contained clinical synopses of variable detail, and were not accompanied by DWI MRI or RT-QuIC results, probably because they predate the advent of these technologies. Finally, co-propagation and contribution of misfolded wild type PrP has previously been demonstrated in P102L, where immunohistochemical discrimination between mutant and wild-type PrP is available.

**Supplementary Figure 2.**
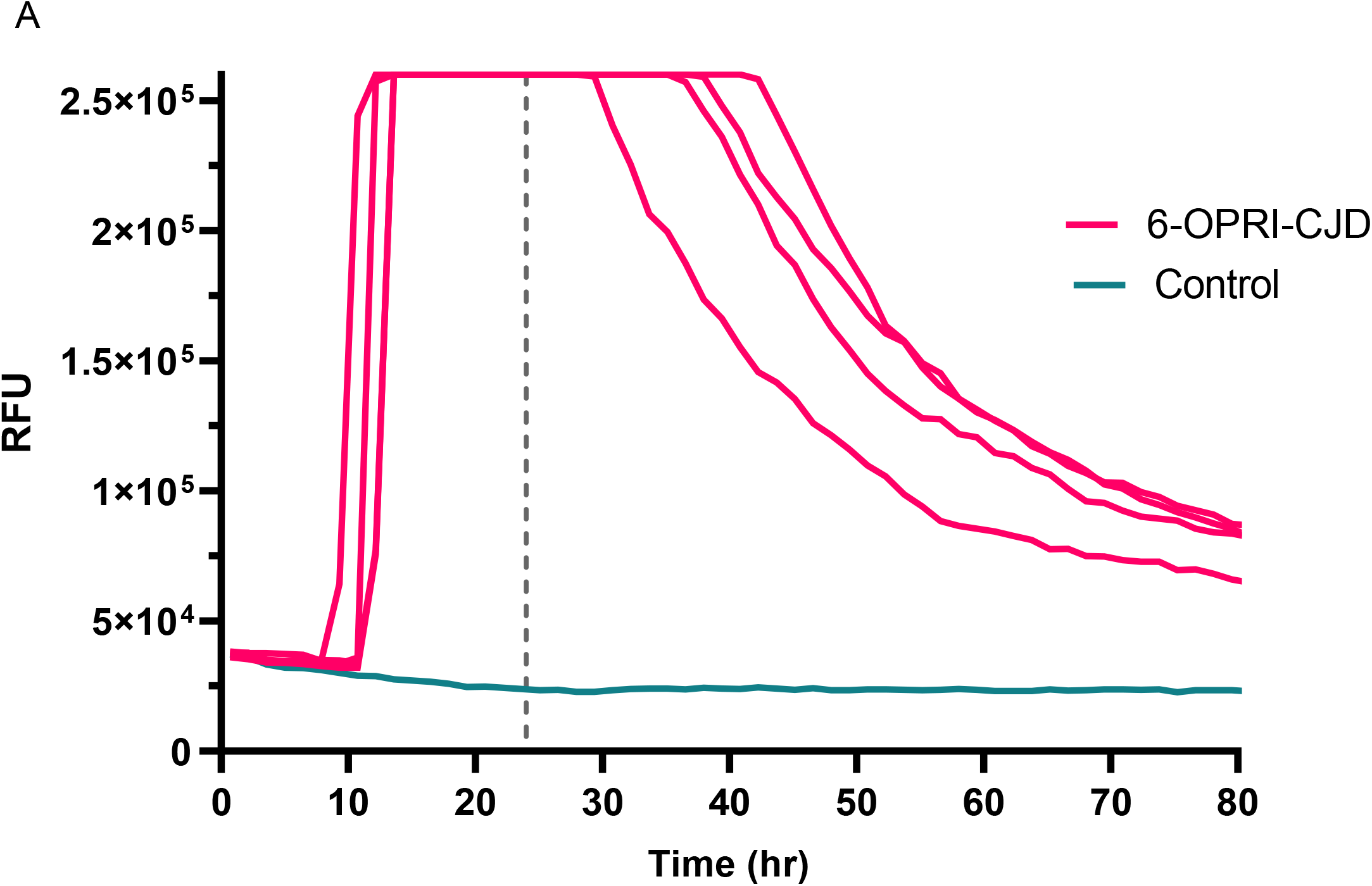
IQ-CSF RT-QuIC graph for 6-OPRI case with “CJD-like” transformation. This graph shows the positive RT-QuIC traces of individual wells against time for this individual with initial classical 6-OPRI phenotype followed by rapid deterioration and death. The dotted vertical line represents the time cut-off for this assay at 24 hours.

#### b. P105S case with positive Hu RT-QuIC with 130 mM NaI at 42°C

These RT-QuIC conditions appear to be able to detect seeding activity from our single symptomatic carrier of P105S who had a CJD-like clinical course of 3 months’ duration; no presymptomatic CSF sample was available to test. The only other P105S case reported in the literature had a completely different clinical course with a disease duration of 10 years; initial MRI Brain showed only cortical atrophy (not clear if DWI was done) but another study 7 years later revealed typical CJD-like DWI changes. This other case predates the clinical application of the CSF RT-QuIC assay. Unlike E200K, it remains to been seen whether this tailored RT-QuIC assay using FL Hu rPrP with NaI is “universal” enough to capture and amplify all P105S related PrP isoforms to be used a diagnostic and/or presymptomatic screening assay.

**Supplementary Figure 3.**
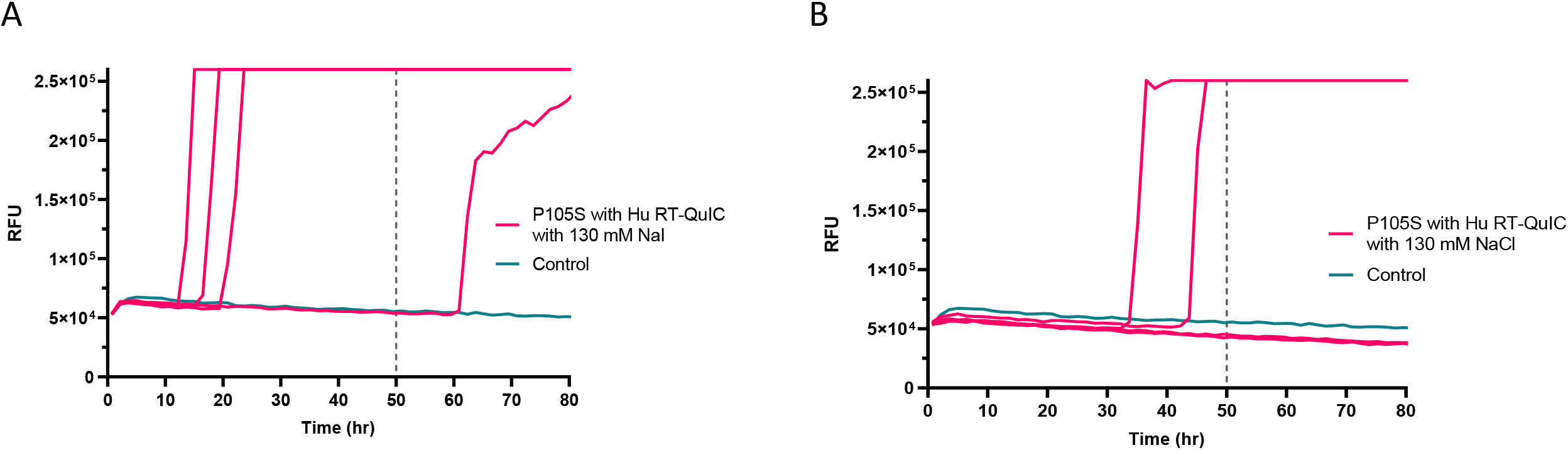
Hu RT-QuIC with 130 mM NaI versus 130 mM NaCl for P105S CSF. (**A**) This shows the RT-QuIC traces of individual wells seeded by CSF from an affected P105S individual, using NaI as salt in the reaction mix. This assay is more sensitive compared to using equivalent NaCl concentration as the salt shown in panel (**B**). The vertical dotted line indicates the time cut-off i.e. 50 hours.

